# Perinatal Impacts of Cannabis and Nicotine: An Analysis of the Cannabis Use During Development and Early Life Study

**DOI:** 10.1101/2024.03.29.24305025

**Authors:** Cassandra J. Trammel, Arpana Agrawal, Ryan Bogdan, Megan Lawlor, Nandini Raghuraman, Jeannie C. Kelly, Christopher D. Smyser, Cynthia Rogers, Ebony B. Carter

**Author notes:** Corresponding author: Cassandra Trammel, MD, MBA: Department of Obstetrics and Gynecology, Washington University School of Medicine, 660 S. Euclid Ave, Box 8064, St. Louis, MO 63110.

## Abstract

**OBJECTIVE:** Evaluate associations between prenatal cannabis use (PCU) and perinatal outcomes.

**METHODS:** We performed an interval analysis of a prospective cohort study of pregnant individuals with pre-pregnancy cannabis use and negative self-report of nicotine use, comparing those who continued cannabis through pregnancy with those who stopped. Patients underwent interviews and urine drug screening for cannabis and cotinine, a nicotine metabolite, in each trimester. The primary outcome was small for gestational age (SGA) at delivery. Secondary outcomes included antenatal and postpartum complications, mode of delivery, and neonatal outcomes. Secondary analyses included stratification by intensity of cannabis use and urinary cotinine positivity.

**RESULTS:** Birthing persons with PCU differed in age (25.5 vs 27.8 years, p=0.001), body mass index (BMI; 27.4 vs 30.9, p=0.001), area deprivation index percentiles (92% PCU vs 88%, p=0.013), cotinine positivity (42.8% vs 10.8%, p<0.001), Hispanic ethnicity (2% vs 7.2%, p=0.009), and education attainment beyond high school (29.4% vs 50%, p<0.001) compared to controls. Birthing person outcomes did not differ. Risks of SGA and other neonatal outcomes did not differ when adjusted for confounders on initial analysis, or with stratification by intensity of cannabis use. Despite negative self-report for nicotine, 42.8% of PCU patients tested positive for cotinine (PCU+c). PCU+c was associated with increased risk of SGA and birthweight less than the 5^th^ percentile, compared to PCU cases without nicotine exposure (17.4% vs 8.3%, aRR 2.7 [1.21-5.38], 35.8% vs 18.6%, aRR 2.4 [1.51-3.48]), and controls (35.8% vs 12.9%, aRR 2.75 [1.64-4.13]). Cotinine-negative PCU patients and controls did not differ.

**CONCLUSION:** PCU was not independently associated with adverse birthing person outcomes. Many patients demonstrated nicotine exposure, either via inadvertent exposure or undisclosed use. While neonates exposed to cannabis alone did not differ from unexposed neonates, those exposed to both cannabis and nicotine were at increased risk of SGA.

## INTRODUCTION

Increasing legalization of medical and recreational cannabis is correlated with rising use, including among pregnant individuals.^1-4^ Large studies and metanalyses have demonstrated potential adverse outcomes associated with prenatal cannabis use (PCU), including anemia and preterm birth.^5,6^ Birthweight is of particular interest; several studies have described an increased risk of low birthweight, and a decreased risk for large for gestational age (LGA) at birth in patients with diabetes who use cannabis relative to diabetic controls.^7-10^ However, concomitant nicotine exposure remains a significant confounder in many studies.^11^ While some prior studies adjust for self-reported tobacco or nicotine use, a majority do not account for inadvertent exposure due to certain preparations of cannabis or secondhand exposure. While retrospective observational studies suggest that outcomes like preterm birth and small for gestational age (SGA, defined as birthweight less than 10^th^ percentile by gestational age) are associated with prenatal cannabis use, these are also well-characterized as a sequelae of prenatal tobacco use.^12-14^

We sought to prospectively evaluate the impact of PCU on outcomes in birthing persons with pre-pregnancy cannabis use and negative self-report of nicotine use, who either continued cannabis in pregnancy or did not. We examined whether PCU, as well as intensity of use, was associated with adverse outcomes for the pregnant person or neonate. Furthermore, we assessed whether associations differed across groups based on positivity for cotinine, a nicotine metabolite, in participants with either inadvertent exposure or potential undisclosed self-administration. We hypothesized that neonates with greater intensity of prenatal cannabis exposure would be at greater risk of SGA, with a possible contribution of concomitant nicotine exposure.

## METHODS

We performed a planned secondary analysis of a single center prospective cohort study of prenatal cannabis exposure beginning July 2019. The Washington University in St. Louis Institutional Review Board (ID 20180001) approved the Cannabis Use During Development and Early Life (CUDDEL) study. All pregnant patients seen in the Barnes-Jewish Hospital Pregnancy clinic were screened for eligibility. Inclusion criteria included pre-pregnancy cannabis use, defined as 1 or more episodes of use prior to knowledge of pregnancy, English proficiency, and ability to provide informed consent. Exclusion criteria included incarceration, age <18 years old, active psychosis, mania, or suicidal ideation, multifetal pregnancy, use of assisted reproductive therapy, self-reported heavy alcohol use, or use of illicit or potentially teratogenic substances (either via self-report or urine screen). Additionally, patients were not eligible for enrollment if they reported tobacco use within the month prior to screening, which occurred during the first trimester of pregnancy.

The overall objectives of the CUDDEL study were to evaluate the impact of PCU on neonatal outcomes, such as SGA, and, in a subset of neonates, to characterize differences in brain and behavioral development related to PCU. Here we present an interim analysis of all patients with neonatal birthweight and gestational age data complete as of 11/7/2023.

Patients, all with a history of cannabis use, were approached in the first trimester of pregnancy. Those who self-reported cannabis use subsequent to knowledge of their pregnancy or had positive urine testing in any trimester were classified as cases (PCU). Those who reported not using cannabis during their pregnancy and tested negative during all trimester visits were classified as controls. Patients were followed with assessments in each trimester and during the delivery admission, which included urine drug screens, self-report of substance use, including both substances and mode of use (e.g., blunts versus joints), and standardized assessments for mental health and stress exposure. Biochemical testing included urine assessment of cotinine, a nicotine metabolite with a detection cutoff 200ng/mL, and 11-Nor-9-carboxy-THC (THCCOOH), a secondary metabolite of cannabis with a detection cutoff of 50ng/mL. Other non-prescribed substances were also evaluated with urinary assays. Biochemical testing results and all self-reports were protected by the Federal Certificate of Confidentiality, stored separately from the medical record, were not disclosed to participants or providers, and thus did not impact clinical care. Assessments and electronic health record review for demographic and outcomes data were completed by trained research staff and compiled in RedCap.

The primary outcome was percent of neonates with SGA at time of delivery. Secondary outcomes for the birthing person included gestational diabetes, anemia, gestational hypertension, preeclampsia, fetal growth restriction, and preterm delivery, as well as mode of delivery, postpartum infections and wound complications, attendance at follow up and breastfeeding. Secondary neonatal outcomes included birthweight less than the 5^th^ percentile for gestational age, large for gestational age at delivery (LGA), 5 minute Apgar score less than 7, and neonatal intensive care unit (NICU) admission. Planned secondary analyses included stratification of outcomes by intensity of cannabis use, defined as “high” (use at least twice monthly in two or more trimesters) or “low” (use at least twice monthly in one trimester). Additional secondary analyses stratified by cotinine positivity were conducted.

Demographic and outcomes data were compared between groups using χ^2^, Mann Whitney U, or Fisher’s exact test for dichotomous variables as appropriate. Relative risks with 95% confidence intervals were calculated for all outcomes. All statistically significant differences between groups were evaluated for possible confounding. A stepwise reverse logistic regression was performed to identify any factors which conferred greater than a 10% change in relative risk, which were considered to be confounders. Confounders were then incorporated in a model to calculate adjusted relative risks. After completing analysis and considering potential collinearity of variables, the final model for birthing person outcomes accounted for obesity, area deprivation index ≥ 75^th^ percentile (ADI, a validated neighborhood-level measure of social exposures, with higher percentiles representing increased socioeconomic disadvantage), and cotinine positivity, while the neonatal outcomes model accounted for ADI ≥ 75^th^ percentile and cotinine.^15^ For secondary analyses comparing cotinine negative and positive groups, cotinine was removed from the adjusted model as groups were already differentiated by the variable of interest. A p-value of <0.05 was considered statistically significant. All statistical analyses were performed using STATA Version 18.0.

## RESULTS

1,023 patients were screened for enrollment. Of 560 eligible patients, 501 were enrolled. At time of analysis, 396 enrolled patients had delivery data available, with 257 in the PCU arm and 139 controls. Patients in the PCU group were significantly younger (age 25.5 vs 27.8 years, p=0.001) and had lower body mass index (BMI) at the initial study visit (27.4 vs 30.9, p=0.001) compared to controls. They were more likely to have a urine drug screen positive for cotinine over the course of the observation (42.8% vs 10.8%, p<0.001), and live in more highly deprived neighborhoods (ADI 92% PCU vs 88% controls, p=0.013), though both groups had high mean ADI. Patients with PCU were less likely to report Hispanic ethnicity (2% vs 7.2%, p=0.009) or educational attainment beyond high school (29.4 vs 50%, p<0.001). There were no significant differences in gestational age at delivery, and most patients in both groups attended three or more prenatal visits.

Birthing person outcomes did not significantly differ between PCU and control groups in unadjusted analysis, but after adjusting for ADI ≥ 75%, obesity (defined as BMI ≥ 30), and cotinine positivity, patients with PCU were more likely to report breastfeeding at discharge (82.1% vs 74.1%, aRR=1.13 [1.01-1.21] (Table 2). The overall postpartum complication rate was too low in both PCU and controls to calculate relative risks for infections or other wound complications. Increased likelihood of SGA less than the 5^th^ and 10^th^ percentiles (12.2% vs 4.32%, RR 2.82 [1.21-6.61] and 26% vs 13%, RR 2.00 [1.24-3.24]) was initially noted for PCU neonates relative to controls, but relative risk estimates were no longer statistically significant (aRR 1.61 [0.63-3.83], aRR 1.32 [0.75-2.20], respectively; Table 3) after adjusting for confounders. Other neonatal outcomes did not differ significantly between groups.

**Table 1:** Demographics and baseline characteristics.

| **Table 1: Demographics and baseline characteristics** |  |  |  |
| --- | --- | --- | --- |
|  | PCU (n= 257) | Controls (n=139) | P value |
| Maternal age in years (median, [range]) | 25.5 [18.8-39.2] | 27.8 [18.7-40.5] | 0.001 |
| Advanced maternal age | 10 (3.9%) | 12 (8.6%) | 0.049 |
| Teen | 21 (8.2%) | 6 (4.3%) | 0.146 |
| Maternal BMI at intake (kg/m2) (median, [range]) | 27.4 [15.5-66.6] | 30.9 [14.8-79.1] | 0.001 |
| Education beyond high school | 70 (29.4%) | 66 (50%) | <0.001 |
| Marital status |  |  | 0.657 |
| Single | 50 (56.8%) | 32 (54.2%) |  |
| Partnered/Married | 36 (40.9%) | 25 (42.4%) |  |
| Divorced/Separated | 1 (1.1%) | 2 (3.4%) |  |
| Widowed | 0 | 0 |  |
| Other* | 1 (1.1%) | 0 |  |
| ADI national percentile (median, [range]) | 92% [38-100%] | 88% [25-100%] | 0.013 |
| ADI >=75%ile | 188 (79.3%) | 86 (69.4%) | 0.035 |
| Race |  |  | 0.06 |
| American Indian/Alaska Native | 2 (0.8%) | 0 (0) |  |
| Asian | 0 (0) | 2 (1.4%) |  |
| Black or African American | 227 (89%) | 113 (81.3%) |  |
| Hawaiian Native & Pacific Islander | 0 (0) | 0 (0) |  |
| White | 25 (9.8%) | 23 (16.6%) |  |
| Other | 1 (0.4%) | 1 (0.7%) |  |
| Ethnicity |  |  | 0.009 |
| Hispanic | 5 (2%) | 10 (7.2%) |  |
| Non-Hispanic | 251 (98%) | 129 (93%) |  |
| Gravity (median, [range]) | 2 [1-6] | 3 [1-6] | 0.104 |
| Prior live births (median, [range]) | 1 [0-5] | 1 [0-5] | 0.756 |
| Concurrent medical conditions |  |  |  |
| Chronic hypertension | 51 (21%) | 36 (27%) | 0.181 |
| Chronic diabetes | 7 (2.9%) | 9 (6.7%) | 0.083 |
| Asthma | 76 (31%) | 42 (30.9%) | 0.957 |
| Anemia | 95 (39.1%) | 51 (37.5%) | 0.760 |
| Psychiatric diagnosis | 123 (50.4%) | 68 (50%) | 0.939 |
| Prior obstetric outcomes |  |  |  |
| Preterm birth | 53 (21.1%) | 24 (17.7%) | 0.415 |
| Hypertensive disorders of pregnancy | 38 (15.6%) | 21 (15.6%) | 0.983 |
| Gestational diabetes | 8 (3.5%) | 5 (4%) | 0.791 |
| Cotinine positivity | 110 (42.8%) | 15 (10.79%) | <0.001 |
| Gestational age at delivery (weeks, [range]) | 38 [24-41] | 38 [23-40] | 0.189 |
| Attendance at 3 or more prenatal visits | 240 (98%) | 136 (99%) | 0.643 |
| Abbreviations: ADI, area of deprivation index; BMI, body mass index; kg, kilograms; m, meters; PCU, prenatal cannabis use |  |  |  |
| *Other indicates that the participant did not self-report a specific race |  |  |  |

**Table 2:**
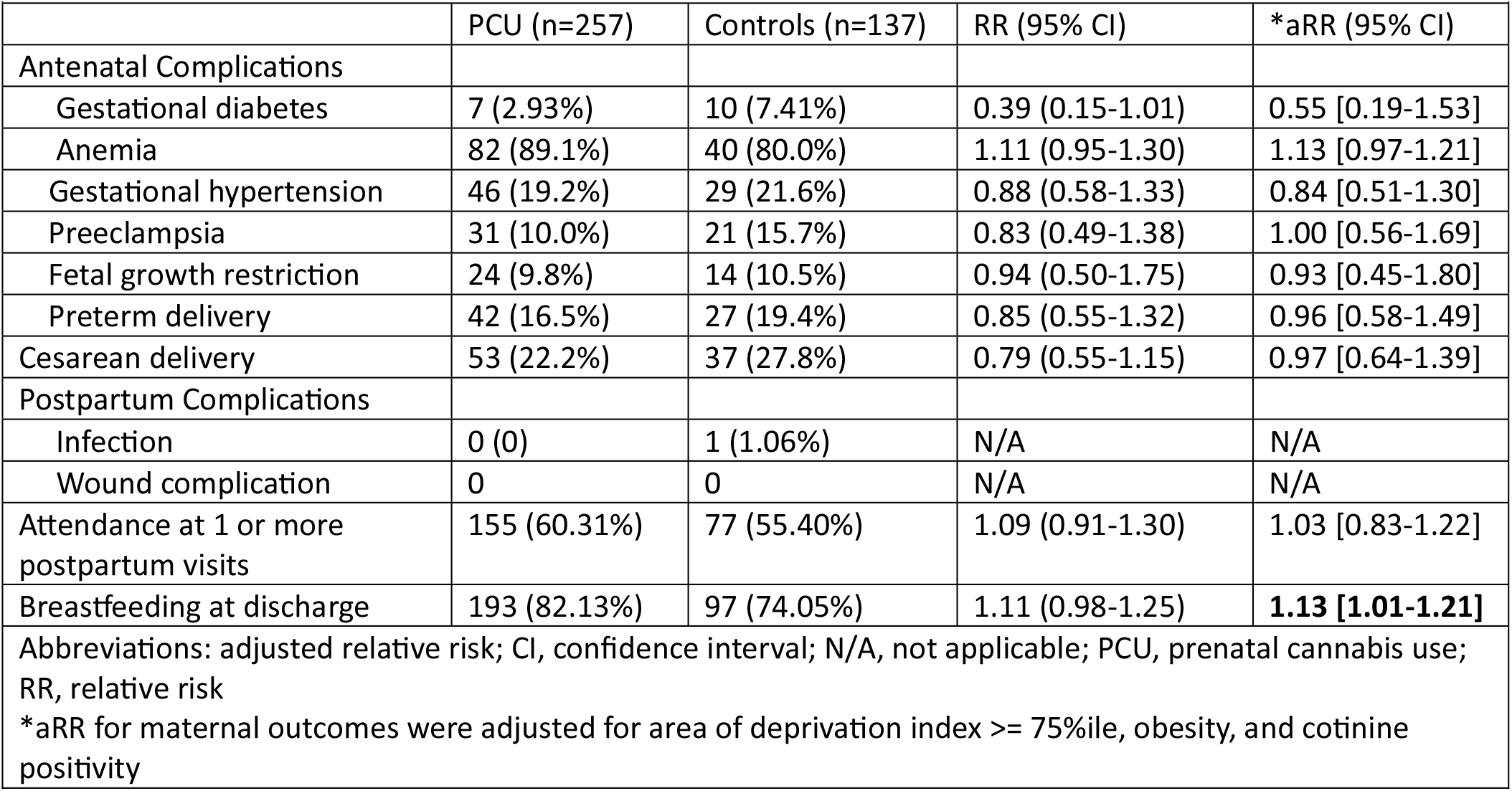
Birthing person outcomes.

**Table 3:** Neonatal outcomes.

| **Table 3: Neonatal outcomes** |  |  |  |  |
| --- | --- | --- | --- | --- |
|  | PCU (n=257) | Controls (n=137) | RR (95% CI) | *aRR (95% CI) |
| SGA < 5%ile | 31 (12.2%) | 6 (4.32%) | 2.82 (1.21-6.61) | 1.61 [0.63-3.83] |
| SGA < 10%ile | 66 (26%) | 18 (13%) | 2.00 (1.24-3.24) | 1.32 [0.75-2.20] |
| LGA | 5 (1.97%) | 8 (5.76%) | 0.34 (0.11-1.02) | 0.45 [0.13-1.46] |
| 5 minute Apgar < 7 | 9 (3.75%) | 9 (6.62%) | 0.56 (0.23-1.39) | 0.50 [0.18-1.32] |
| NICU admission | 43 (17.55%) | 24 (17.65%) | 0.99 (0.63-1.56) | 0.87 [0.50-1.44] |
| Abbreviations: aRR, adjusted relative risk ; CI, confidence interval; N/A, not applicable; LGA, large for gestational age; NICU, neonatal intensive care unit; PCU, prenatal cannabis use; RR, relative risk; SGA, small for gestational age |  |  |  |  |
| *aRR for neonatal outcomes were adjusted for area of deprivation index >= 75%ile and cotinine positivity |  |  |  |  |

On secondary analysis stratifying by PCU use at least twice monthly in ≥ 2 trimesters (high, PCU-H, n=77) versus 1 trimester (low, PCU-L, n=180), PCU-H patients differed from controls in age, BMI, educational attainment, and cotinine positivity (Appendix, Table 4). While there were no statistically significant differences in birthing person outcomes between PCU-H patients and controls (Appendix, Table 5), PCU-neonates had a higher likelihood of SGA less than the 5^th^ and 10^th^ percentiles than controls (13.2% vs 4.32%, RR 3.05 [1.15-8.06], 30.3% vs 13%, RR 2.34 [1.35-4.05]) on unadjusted analysis (Figure 1). Low PCU (PCU-L) neonates also had higher risk of SGA on unadjusted analysis relative to controls (11.8% vs 4.3%, RR 2.73 [1.13-6.59], 24.2% vs 13%, RR 1.87 [1.12-3.09]), with no significant difference between PCU-H and PCU-L patients (Figure 1). However, adjusting for confounders rendered the relative risks for SGA statistically non-significant (PCU-H vs controls: aRR 1.44 [0.39-4.68] and aRR 1.53 [0.72-2.88]; PCU-L vs controls aRR 1.8 [0.69-4.31] and aRR 1.31 [0.71-2.24]) (Figure 1).

**Figure 1:**
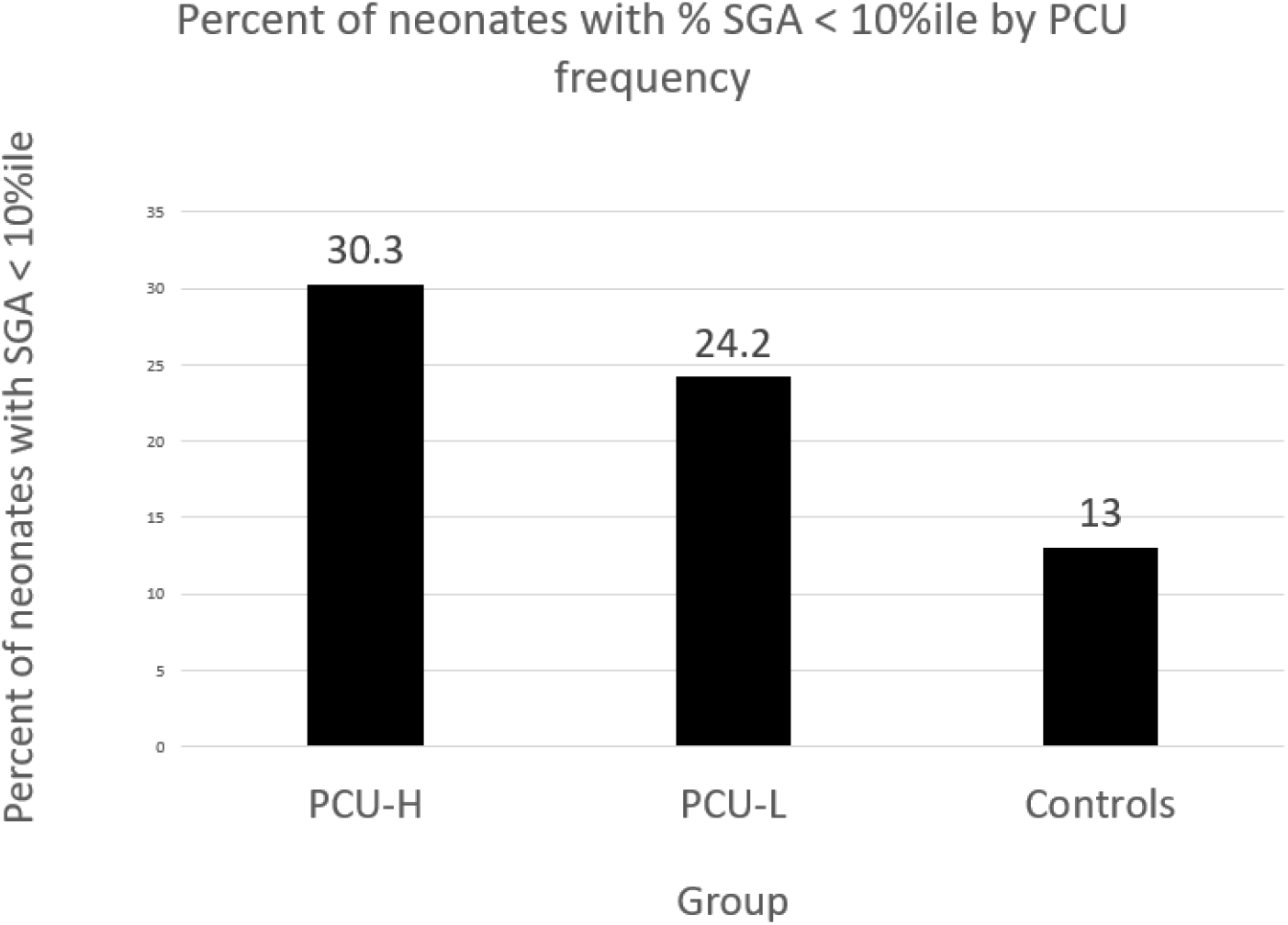
Percentage of SGA neonates by PCU frequency. On initial analysis, patients who reported more high frequency cannabis use had significantly higher rates of SGA than controls (RR 2.34, 95% CI [1.35‐ 4.05]), and patients with low frequency use also had higher rates of SGA neonates (RR 1.87 95% CI [1.12‐ 3.09]), while there were no differences between the PCU‐H and L groups. However, on adjusted analysis, there was no statistically significant difference between PCU‐H and controls (aRR 1.52, 95% CI [0.72‐2.88]) or PCU‐L and controls (aRR1.31 95% CI [0.71‐2.24]). Abbreviations: aRR, adjusted relative risk; PCU, prenatal cannabis use; PCU‐H, high frequency PCU use; PCU‐L, low frequency PCU use; RR, relative risk; SGA, small for gestational age

For the purposes of secondary analysis by nicotine exposure, the 15 controls who tested positive for cotinine were excluded from the control group. When stratifying by cotinine positivity, PCU patients with cotinine (PCU+c; n=110) significantly differed from PCU patients without cotinine (PCU-c; n=147) with regard to ADI percentile (94% vs 89%, p=0.017) and history of a hypertensive disorder of pregnancy in a prior pregnancy (22.1% vs 10.8%, p=0.016) (Appendix, Table 7). Although PCU+c patients were more likely to attend a postpartum visit than PCU-c patients (68.2 vs 54.4%, RR 1.25 [1.03-1.52], aRR 1.25 [1.0047-1.45]) (Appendix, Table 8), the remainder of birthing person outcomes did not significantly differ across these groups. PCU+c patients were significantly more likely to have a neonate with SGA less than the 5^th^ and 10^th^ percentiles compared to PCU-c patients (17.4% vs 8.3%, RR 2.1 [1.07-4.15], 35.8% vs 18.5%, RR 1.92 [1.26-2.93]); this difference persisted after controlling for confounders (aRR 2.7 [1.21-5.38], aRR 2.4 [1.51-3.48] (Figure 2). Similarly, when comparing PCU+c cases to cotinine-negative controls, the increased risk of SGA less than the 5^th^ and 10^th^ percentiles persisted after adjustment for ADI (17.4% vs 4.8%, aRR 3.29 [1.33-6.95], 35.8% vs 12.9%, aRR 2.75 [1.64-4.13]. However there was not a statistically significant difference in risk of SGA between PCU-c cases and controls (Figure 2).

**Figure 2:**
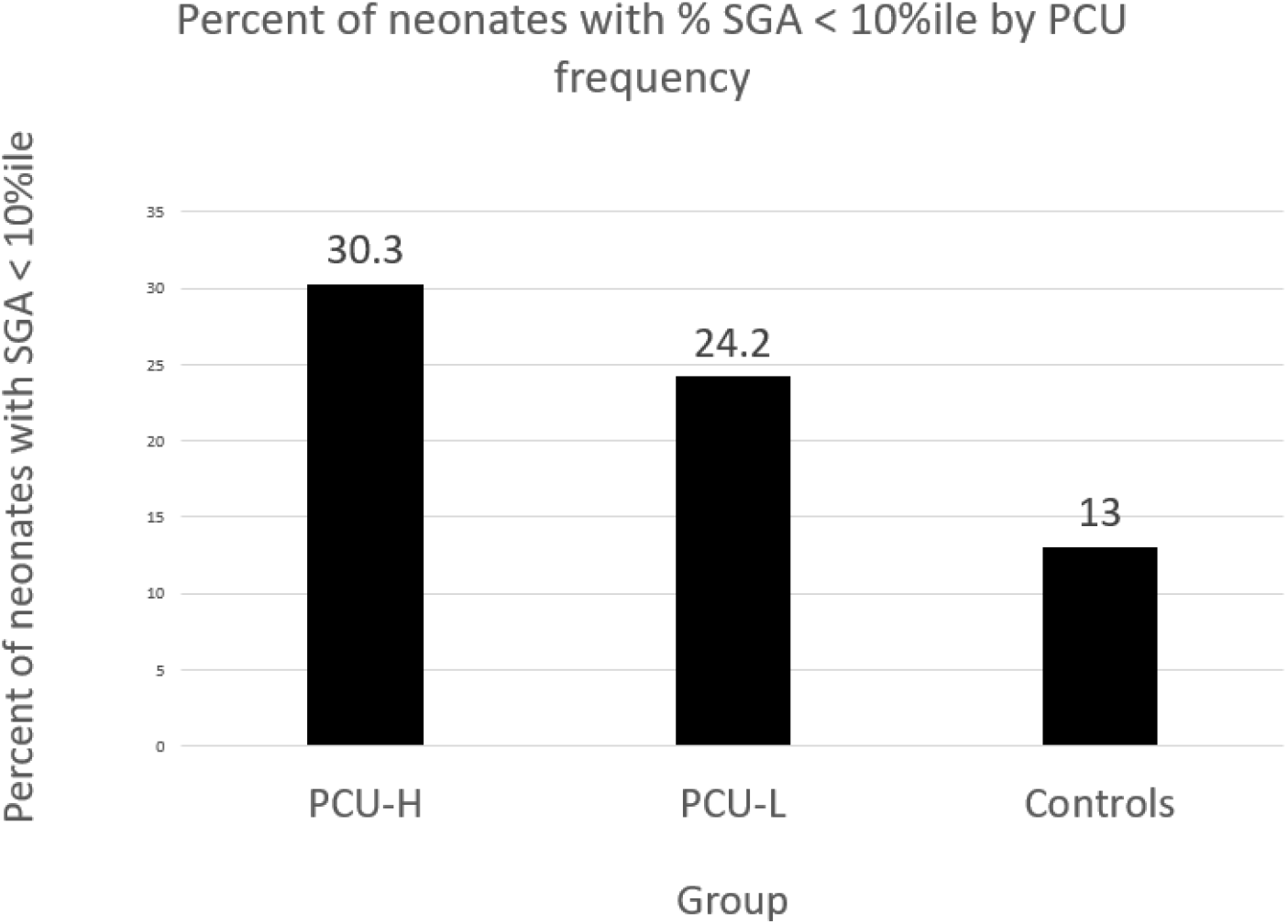
Percentage of SGA neonates by PCU frequency. On initial analysis, patients who reported more high frequency cannabis use had significantly higher rates of SGA than controls (RR 2.34, 95% CI [1.35‐ 4.05]), and patients with low frequency use also had higher rates of SGA neonates (RR 1.87 95% CI [1.12‐ 3.09]), while there were no differences between the PCU‐H and L groups. However, on adjusted analysis, there was no statistically significant difference between PCU‐H and controls (aRR 1.52, 95% CI [0.72‐2.88]) or PCU‐L and controls (aRR1.31 95% CI [0.71‐2.24]). Abbreviations: aRR, adjusted relative risk; PCU, prenatal cannabis use; PCU‐H, high frequency PCU use; PCU‐L, low frequency PCU use; RR, relative risk; SGA, small for gestational age

## DISCUSSION

Our study found that prenatal cannabis use was not significantly associated with differences in major outcomes for the birthing persons, including antenatal diagnoses and mode of delivery, across subgroup analyses. These findings align with existing data that cannabis use in pregnancy does not appear to significantly impact birthing person outcomes, or that effect sizes associated with these outcomes are too modest to be detected in the current cohort. The only difference in birthing person outcomes that persisted after adjustment was breastfeeding at discharge, with PCU patients more likely to breastfeed than controls. On subgroup analysis, attendance at postpartum visits differed, with PCU+c patients significantly more likely to follow up postpartum than their PCU-c counterparts. As conditions requiring closer follow up such as gestational hypertension or cesarean delivery did not differ between groups, the mechanism underlying this finding is unclear. Both groups were similar in likelihood of attending a follow-up visit (68% in PCU+c, and 54% in PCU-c) to the 60% follow up rate reported by ACOG for all postpartum patients (ACOG optimizing pp care). Additionally, secondary neonatal outcomes did not differ across groups. While we have previously described lower rates of LGA among patients with diabetes who use cannabis at our institution, we did not find significant differences between gestational diabetes or LGA relative to cannabis use in this study.

For our primary outcome of interest, SGA, initial analyses indicated a higher risk of SGA in PCU patients, particularly in those using cannabis at least twice a month for two or more trimesters, consistent with some prior studies.^16^ However, after adjusting for confounders, including cotinine, there were no statistically significant differences in neonatal outcomes between PCU and control patients, which has also been previously demonstrated in the literature.^17^ It is possible that this finding represents a type 2 error related to sample size, since the point estimate of our aRR (Table 3; aRR=1.61 [95% C.I. 0.63 – 3.83]) was highly comparable to the aRR for the same outcome in a recent study (aRR 1.52 [95% C.I. 1.08 – 2.14]) with a larger sample size.^16^ Similarly, when comparing SGA rates between high PCU, low PCU, and controls, there were no significant differences once accounting for ADI. In contrast, analyses comparing SGA across PCU+c and controls demonstrated that neonates exposed to both cannabis and nicotine had significantly higher risk of SGA than both those exposed to cannabis alone and to controls, with no difference between PCU-c neonates and controls.

Because self-report of tobacco use was a reason for exclusion, we hypothesize most cotinine positivity, and therefore, nicotine exposure, was incidental and may be attributable to the route of cannabis administration; in our cohort, we have previously found that 75% of patients with both self-report and urine positivity for cannabis use used blunts. Blunts are made by hollowing out a cigar and filling it with cannabis, and may therefore include tobacco, contrasted with typical joints or smokeless products such as edibles or vaporizers. Given the high rate of blunt use in our cohort, any associations with cotinine in the PCU group (i.e., PCU+c) may be confounded by intensity of cannabis use, as patients with tobacco contaminant in their cannabis would be more likely to have cotinine positivity if they were using cannabis more frequently; indeed, there was greater cotinine positivity in PCU-H vs. PCU-L (54.6% vs 10.8%, p<0.001) and a greater proportion of high-frequency cannabis use in PCU+c vs. PCU-c (38.18% vs 23.81%, p=0.013).

Our study expands the existing body of literature by prospectively assessing degree of cannabis use in pregnancy, including mode of use, and by excluding patients with concomitant disclosed nicotine use. Additionally, we recruited only patients with a history of cannabis use prior to pregnancy for both arms of the study, which provides effective statistical control for factors associated with any lifetime cannabis use. Strengths of the current study include the prospective collection of data, which may reduce recall bias, as well as incorporation of both patient report of frequency and modes of cannabis use, urine testing of all patients in each trimester, and urine assessment of tobacco exposure.

Potential limitations include high rates of cotinine positivity and its prominent confounding effect; because of the need to control for cotinine in adjusted analyses, this may have increased the likelihood of a type 2 error specific to cannabis-only exposures. Because we excluded participants who self-reported tobacco use, few controls tested positive for cotinine. Thus, we were unable to evaluate any potential synergistic effect of cannabis and nicotine relative to either substance alone. Additionally, due to high research recruitment in our patient population and a concurrent study regarding diabetes prevention in pregnancy within the same clinic, GDM rates are low in our PCU group, which may impact LGA findings. Finally, findings in this analysis are limited to birthing person and neonatal outcomes surrounding the birthing event, and do not include long-term psychobehavioral sequelae of exposure that have been described in other studies; the larger ongoing CUDDEL study will follow these neonates into childhood.

For future studies, these findings further underscore the importance of evaluating concomitant nicotine exposure in studies of prenatal cannabis use for research purposes. Particularly in cohorts where blunt usage is common, such as ours, exclusion based on self-report of tobacco use may not be feasible, and biological cotinine testing should be considered for research purposes. However, prospective exclusion of patients based on mode of cannabis use or risk of cotinine exposure may disproportionately exclude populations in which blunts are commonplace and thus, might be inadvisable from a research equity perspective. Additionally, our findings regarding differential rates of breastfeeding and follow up, while not clinically actionable, could both feasibly impact childhood outcomes. Comprehensive data collection to assess for potential confounders will be important in interpreting childhood outcomes in future studies.

Clinically, our finding of high rates of inadvertent or undisclosed nicotine exposure may be helpful in counseling patients who opt to continue cannabis use during pregnancy. Educating patients regarding the possible presence of tobacco in products used for consumption of cannabis, such as blunts and certain joint rolling papers, with potential risks regarding neonatal growth, is critical. However, in the absence of conclusive findings regarding significant perinatal complications for the birthing person or neonate associated with urine-proven cannabis use, our findings also underscore the importance of thoughtful consideration regarding the utility of including cannabis on clinical (i.e., not related to research) urine screens, as but such tests may result in significant legal and emotional ramifications for parenting people.^18^

While isolated cannabis exposure did not appear to significantly impact parental or neonatal outcomes in our cohort, our findings emphasize the importance of careful history taking and counseling regarding potential risks. PCU may impact several parental variables not evaluated in this study, such as psychologic well-being, admissions for nausea and vomiting, adequate prenatal weight gain, and postpartum depression. Our finding of increased likelihood of SGA in birthing persons who were concomitantly exposed to nicotine, and were also heavier cannabis users, urges caution regarding the use of cannabis and tobacco during the prenatal period, consistent with other large studies and recommendations of the ACOG, as developmental sequelae of such exposures as the child ages remain understudied. Future directions of the CUDDEL project include ongoing multimodal assessment of neonates into childhood to help further clarify these risks.

## Data Availability

All data produced in the present study are currently not available upon request as the larger study is ongoing.

