## Appendices for "Perinatal Impacts of Cannabis and Nicotine: An Analysis of the Cannabis Use During Development and Early Life Study"

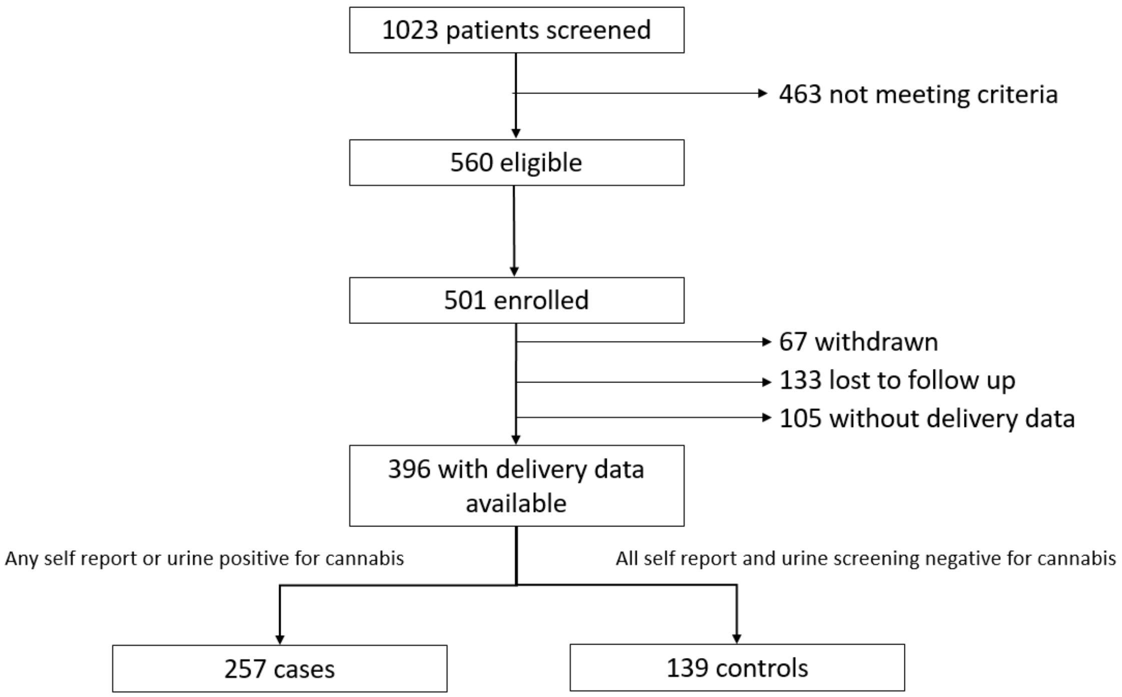

| **Table 4: Demographics by PCU intensity (PCU-H vs Controls)** | | | |
|---|---|---|---|
| | PCU-H (N = 77) | Controls (N = 137) | P value |
| Maternal age (years) | 25.6 [18.8-37.4] | 27.8 [18.7-40.5] | 0.0033 |
| AMA | 1 (1.3%) | 12 (8.63%) | 0.03 |
| Teen | 8 (10.4%) | 6 (4.32%) | 0.082 |
| Maternal BMI at intake (kg/m2) | 28.7 [17.1-57.3] | 33 [14.8-79.1] | 0.0049 |
| Education beyond high school | 26 (33.8%) | 66 (50%) | 0.023 |
| Marital status | | | 0.495 |
| Single | 20 (60.6%) | 32 (54.2%) | |
| Partnered/Married | 11 (33.3%) | 25 (42.4%) | |
| Divorced/Separated | 1 (3%) | 2 (3.4%) | |
| Widowed | 0 | 0 | |
| Other | 1 (3%) | 0 | |
| ADI (national) | 81.4 [31-100] | 79.3 [17-100] | 0.66 |
| ADI >=75%ile | | | 0.490 |
| Race | | | 0.59 |
| Black or African American | 66 (85.7%) | 113 (81.3%) | |
| White | 11 (14.3%) | 23 (16.6%) | |
| American Indian/Alaska N | 0 | 0 | |
| Asian | 0 | 2 (1.4%) | |
| Hawaiian N. & Pacific Is. | 0 | 0 | |
| Other | 0 | 1 (0.7%) | |
| Ethnicity | | | 0.059 |

| | | | |
|---|---|---|---|
| Hispanic | 1 (1.3%) | 10 (7.2%) | |
| Non-Hispanic | 76 (98.7%) | 129 (92.8%) | |
| Gravity (median, [range]) | 2.7 [1-6] | 3 [1-6] | 0.4183 |
| Prior live births (median, [range]) | 1 [0-4] | 1 [0-5] | 0.8915 |
| Concurrent medical conditions | | | |
| Chronic hypertension | 14 (18.4%) | 36 (27.1%) | 0.159 |
| Chronic diabetes | 1 (1.45%) | 9 (6.7%) | 0.083 |
| Asthma | 21 (27.6%) | 42 (30.9%) | 0.619 |
| Anemia | 25 (33.3%) | 51 (37.5%) | 0.546 |
| Psychiatric diagnosis | 32 (42.1%) | 68 (50%) | 0.269 |
| Prior obstetric outcomes | | | |
| Preterm birth | 15 (19.7%) | 24 (17.7%) | 0.706 |
| Hypertensive disorders of pregnancy | 17 (22.4%) | 21 (15.6%) | 0.216 |
| Gestational diabetes | 2 (2.7%) | 5 (4%) | 0.643 |
| Cotinine positivity | 42 (54.6%) | 15 (10.8%) | <0.00 |
| Gestational age at delivery (weeks) | 37.5 [27-41] | 37.5 [23-40] | 0.6370 |
| Attendance at 3 or more prenatal visits | 75 (99.7%) | 136 (99.3%) | 0.671 |

Abbreviations: ADI, area of deprivation index; BMI, body mass index; kg, kilograms; m, meters; PCU-H, high prenatal cannabis use
*Other indicates that the participant did not self-report a specific race

**Table 5: Birthing person outcomes by PCU intensity (PCU-H vs Controls)**

| | PCU-H (n=77) | Controls (n=139) | RR (95% CI) | *aRR (95% CI) |
|---|---|---|---|---|
| Antenatal Complications | | | | |
| Gestational diabetes | 0 (0%) | 10 (8.0%) | N/A | N/A |
| Anemia | 21 (87.5%) | 40 (80%) | 1.09 [0.89-1.34] | 1.06 [0.68-1.20] |
| Gestational hypertension | 19 (25%) | 29 (21.6%) | 1.16 [0.7-1.91] | 1.05 [0.54-1.82] |
| Preeclampsia | 11 (14.7%) | 21 (15.7%) | 0.93 [0.48-1.84] | 1.19 [0.52-2.39] |
| Fetal growth restriction | 8 (11%) | 14 (10.5%) | 1.04 [0.46-2.38] | 1.22 [0.44-2.98] |
| Preterm delivery | 17 (22.4%) | 27 (19.4%) | 1.15 [0.67-1.97] | 1.54 [0.81-2.54] |
| Cesarean delivery | 15 (20%) | 37 (27.8%) | 0.72 [0.42-1.22] | 0.84 [0.43-1.46] |
| Breastfeeding at discharge | 60 (80%) | 97 (74.1%) | 1.08 [0.93-1.28] | 1.08 [0.87-1.21] |
| Attendance at 1 or more postpartum visits | 42 (54.6%) | 77 (55.4%) | 0.98 [0.76-1.27] | 0.92 [0.63-1.21] |

Abbreviations: adjusted relative risk; CI, confidence interval; N/A, not applicable; PCU-H, high prenatal cannabis use; RR, relative risk
*aRR for maternal outcomes were adjusted for area of deprivation index >= 75%ile, obesity, and cotinine positivity

**Table 6: Neonatal outcomes by PCU intensity**

| | PCU-H (n=77) | Controls (n=139) | RR (95% CI) | *aRR (95% CI) |
|---|---|---|---|---|
| SGA < 5ile | 10 (13.2%) | 6 (4.32%) | **3.05 [1.15-8.06]** | 1.44 [0.39-4.68] |
| SGA < 10ile | 23 (30.3%) | 18 (13%) | **2.34 [1.35-4.05]** | 1.53 [0.72-2.88] |
| LGA | 1 (1.3%) | 8 (5.8%) | 0.23 [0.03-1.79] | 0.16 [0.02-1.59] |
| 5 min Agpar < 7 | 1 (1.3%) | 9 (6.6%) | 0.2 [0.03-1.54] | 0.15 [0.02-1.34] |

| | | | | |
|---|---|---|---|---|
| NICU admission | 11 (14.9%) | 24 (17.7%) | 0.84 [0.44-1.62] | 0.81 [0.35-1.71] |
| | PCU-H (n=77) | PCU-L (n=180) | RR (95% CI) | *aRR (95% CI) |
| SGA < 5ile | 10 (13.2%) | 21 (11.8%) | 1.11 [0.55-2.25] | 0.73 [0.29-1.71] |
| SGA < 10ile | 23 (30.3%) | 43 (24.2%) | 1.25 [0.81-1.92] | 1.05 [0.6-1.69] |
| LGA | 1 (1.3%) | 4 (2.3%) | 0.59 {0.07-5.15] | 0.77 [0.08-6.27] |
| 5 min Agpar < 7 | 1 (1.3%) | 8 (4.9%) | 0.27 [0.03-2.12] | 0.28 [0.03-2.12] |
| NICU admission | 11 (14.9%) | 32 (18.7%) | 0.79 [0.42-1.49] | 0.82 [0.39-1.61] |
| | PCU-L (n=180) | Controls (n=139) | RR (95% CI) | *aRR (95% CI) |
| SGA < 5ile | 21 (11.8%) | 6 (4.3%) | **2.73 [1.13-6.59]** | 1.8 [0.69-4.31] |
| SGA < 10ile | 43 (24.2%) | 18 (13%) | **1.87 [1.12-3.09]** | 1.31 [0.71-2.24] |
| LGA | 4 (2.3%) | 8 (5.8%) | 0.39 [0.12-1.27] | 0.45 [0.12-1.59] |
| 5 min Agpar < 7 | 8 (4.9%) | 9 (6.6%) | 0.74 [0.29-1.86] | 0.66 [0.24-1.71] |
| NICU admission | 32 (18.7%) | 24 (17.7%) | 1.06 [0.6601.71] | 0.97 [0.5-1.74] |

Abbreviations: aRR, adjusted relative risk ; CI, confidence interval; N/A, not applicable; LGA, large for gestational age; NICU, neonatal intensive care unit; PCU-H, high prenatal cannabis use; PCU-L, low prenatal cannabis use; RR, relative risk; SGA, small for gestational age
*aRR for neonatal outcomes were adjusted for area of deprivation index >= 75%ile and cotinine positivity

**Table 7: Demographics by cotinine positivity**

| | PCU+c (N= 110) | PCU-c (N= 147) | P value |
|---|---|---|---|
| Maternal age in years (median, [range]) | 24.9 [18.8-38.6] | 26.1 [19.1-39.2] | 0.093 |
| AMA | 3 (2.7%) | 7 (4.8%) | 0.404 |
| Teen | 10 (9.1%) | 11 (7.5%) | 0.641 |
| Maternal BMI at intake (kg/m2) (median, [range]) | 28.2 [15.5-66.6] | 27.1 [15.8-58.1] | 0.929 |
| Education beyond high school | 26 (25.5%) | 44 (32.4%) | 0.25 |
| Marital status | | | 0.208 |
| Single | 20 (66.7%) | 30 (51.7%) | |
| Partnered/Married | 9 (30%) | 27 (46.6%) | |
| Divorced/Separated | 1 (3.3%) | 0 | |
| Widowed | 0 | 0 | |
| Other | 0 | 1 (1.7%) | |
| ADI (national) | 94 [31-100] | 89 [34-100] | 0.017 |
| ADI >=75%ile | 88 (86.3%) | 100 (74.1%) | 0.022 |
| Race | | | 0.413 |
| Black or African American | 100 (91.7%) | 127 (87%) | |
| White | 9 (8.3%) | 16 (11%) | |
| American Indian/Alaska N | 0 | 2 (1.4%) | |
| Asian | 0 | 0 | |
| Hawaiian N. & Pacific Is. | 0 | 0 | |
| Other | 0 | 1 (0.7%) | |
| Ethnicity | | | |
| Hispanic | 2 (1.8%) | 3 (2.0%) | |

| | | | |
|---|---|---|---|
| Non-Hispanic | 107 (98.2%) | 144 (98%) | |
| Gravity (median, [range]) | 2 (1-6) | 2 (1-6) | 0.971 |
| Prior live births (median, [range]) | 1 (0-5) | 1 (0-5) | 0.677 |
| Concurrent medical conditions | | | |
| Chronic hypertension | 21 (20.2%) | 30 21.6%) | 0.792 |
| Chronic diabetes | 2 (1.9%) | 5 (3.7%) | 0.406 |
| Asthma | 33 (31.4%) | 43 (31%) | 0.934 |
| Anemia | 39 (37.1%) | 56 (40.6%) | 0.587 |
| Psychiatric diagnosis | 55 (52.4%) | 68 (49%) | 0.592 |
| Prior obstetric outcomes | | | |
| Preterm birth | 25 (23.4%) | 28 (19.4%) | 0.452 |
| Hypertensive disorders of pregnancy | 23 (22.1%) | 15 (10.8%) | 0.016 |
| Gestational diabetes | 4 (3.9%) | 4 (3.1%) | 0.745 |
| Gestational age at delivery (weeks) | 39 [24-40] | 38 [27-41] | 0.608 |
| Attendance at 3 or more prenatal visits | 104 (99.1%) | 136 (98.6%) | 0.728 |

Abbreviations: ADI, area of deprivation index; BMI, body mass index; kg, kilograms; m, meters; PCU+c, prenatal cannabis use with cotinine positivity; PCU-c, prenatal cannabis use with negative cotinine
*Other indicates that the participant did not self-report a specific race

**Table 8: Birthing person outcomes among PCU patients by cotinine status**

| | PCU+cot (n=110) | PCU-cot (n=147) | RR (95% CI) | *aRR (95% CI) |
|---|---|---|---|---|
| Antenatal Complications | | | | |
| Gestational diabetes | 1 (1.0%) | 6 (4.4%) | 0.22 [0.02-1.8] | 0.21 [0.02-1.78] |
| Anemia | 32 (88.9%) | 50 (89.3%) | 1.0 [0.86-1.15] | 0.93 [0.6-1.06] |
| Gestational hypertension | 24 (23.3%) | 22 (16.1%) | 1.45 [0.86-1.44] | 1.44 [0.83-2.31] |
| Preeclampsia | 10 (9.8%) | 21 (15.4%) | 0.63 [0.31-1.29] | 0.66 [0.30-1.33] |
| Fetal growth restriction | 11 (10.5%) | 13 (9.4%) | 1.12 [0.52-2.4] | 1.16 [0.49-2.51] |
| Preterm delivery | 16 (14.7%) | 26 (17.9%) | 0.82 [0.46-1.45] | 0.83 [0.43-1.52] |
| Cesarean delivery | 20 (19.4%) | 33 (24.3%) | 0.80 [0.49-1.31] | 0.81 [0.46-1.35] |
| Breastfeeding at discharge | 87 (83.7%) | 106 (80.9%) | 1.03 [0.92-1.16] | 1.03 [0.88-1.13] |
| Attendance at 1 or more postpartum visits | 75 (68.2%) | 80 (54.4%) | **1.25 [1.03-1.52]** | **1.25 [1.00-1.45]** |

Abbreviations: adjusted relative risk; CI, confidence interval; N/A, not applicable; PCU+c, prenatal cannabis use with cotinine positivity; PCU-c, prenatal cannabis use with negative cotinine; RR, relative risk
*aRR for maternal outcomes were adjusted for area of deprivation index >= 75%ile and obesity

**Table 9: Neonatal outcomes by cotinine status**

| | PCU+cot (n=110) | PCU-cot (n=147) | RR (95% CI) | *aRR (95% CI) |
|---|---|---|---|---|
| SGA < 5ile | 19 (17.4%) | 12 (8.3%) | **2.1 [1.07-4.15]** | **2.7 [1.21-5.38]** |
| SGA < 10ile | 39 (35.8%) | 27 (18.6%) | **1.92 [1.26-2.93]** | **2.4 [1.51-3.48]** |
| LGA | 0 (0) | 5 (3.5%) | N/A | N/A |
| 5 min Agpar < 7 | 4 (3.9%) | 5 (3.6%) | 1.08 [0.3-3.93] | 0.94 [0.25-3.27] |
| NICU admission | 18 (17.4%) | 25 (17.9%) | 0.96 [0.55-1.66] | 0.84 [0.43-1.53] |

| | | | | |
|---|---|---|---|---|
| | PCU+cot (n=110) | Controls (n=124) | RR (95% CI) | *aRR (95% CI) |
| SGA < 5ile | 19 (17.4%) | 6 (4.8%) | **3.60 [1.50-8.69]** | **3.29 [1.33-6.95]** |
| SGA < 10ile | 39 (35.8%) | 16 (12.9%) | **2.77 [1.64-4.67]** | **2.75 [1.64-4.13]** |
| LGA | 0 (0%) | 6 (4.8%) | Unable to calc | Unable to calc |
| 5 min Agpar < 7 | 4 (3.9%) | 8 (6.6%) | 0.59 [0.18-1.91] | 0.53 [0.16-1.69] |
| NICU admission | 18 (17.1%) | 22 (18.2%) | 0.94 [0.54-1.66] | 0.78 [0.4-1.44] |
| | PCU-cot (n=110) | Controls (n=124) | RR (95% CI) | *aRR (95% CI) |
| SGA < 5ile | 12 (8.3%) | 6 (4.8%) | 1.71 [0.66-4.42] | 1.15 [0.40-3.05] |
| SGA < 10ile | 27 (18.6%) | 16 (12.9%) | 1.44 [0.82-2.55] | 1.15 [0.59-2.08] |
| LGA | 5 (3.5%) | 6 (4.8%) | 0.71 [0.22-2.28] | 0.87 [0.25-2.81] |
| 5 min Agpar < 7 | 8 (6.6%) | 5 (3.6%) | 0.55 [0.18-1.63] | 0.51 [0.16-1.49] |
| NICU admission | 22 (18.2%) | 25 (17.9%) | 0.98 [0.58-1.65] | 0.87-0.48] |

Abbreviations: aRR, adjusted relative risk ; CI, confidence interval; N/A, not applicable; LGA, large for gestational age; NICU, neonatal intensive care unit; PCU+c, prenatal cannabis use with cotinine positivity; PCU-c, prenatal cannabis use with negative cotinine; RR, relative risk; SGA, small for gestational age

*aRR for neonatal outcomes were adjusted for area of deprivation index >= 75%ile
